# Association of long-term body weight variability with dementia: a prospective study

**DOI:** 10.1101/2021.07.19.21258665

**Authors:** Hui Chen, Tianjing Zhou, Jie Guo, John S. Ji, Liyan Huang, Weili Xu, Guangmin Zuo, Xiaozhen Lv, Yan Zheng, Albert Hofman, Yuan Ma, Changzheng Yuan

## Abstract

**Introduction**

We aimed to investigate whether long-term body weight variability (BWV) is associated with late-life dementia and to further assess their potential temporal relationships.

**Methods**

In 5,547 participants in Health and Retirement Study (HRS), a population-based prospective cohort, we quantified BWV as coefficient of variation using self-reported body weight from 1992 to 2008 and followed their dementia status from 2008 to 2016.

**Results**

A total of 427 incident dementia cases were identified. Larger long-term BWV was significantly associated with higher risk of dementia (HR comparing extreme quartiles: 2.01, 95% CI: 1.48-2.72; HR of each SD increment: 1.21, 95% CI,1.10-1.32; p-trend<0.001). This significant association was even observed for BWV estimated approximately 15 years preceding dementia diagnosis (HR of each SD increment: 1.13, 95% CI: 1.03-1.23) and was more pronounced for that closer to diagnosis.

**Discussion**

Our findings suggested that large BWV could be a novel risk factor for dementia.

## 1. INTRODUCTION

Dementia, a major type of neurodegenerative disease, poses tremendous burdens on older people’s well-being and caring cost worldwide[1]. Body weight and its long-term change have been linked with dementia in later life[2–6], but the overall relationship is still not fully understood[2, 7–8]. Body weight variability (BWV) measures the intraindividual instability and fluctuations in body weight during a period. Unlike static body weight and body weight change, BWV mainly captures the instability of body weight over time and has been associated with higher risks of multiple morbidities and mortality[9–13]. Recently, emerging evidence has also suggested that larger body weight fluctuation was associated with a higher risk of dementia[14–16]. However, the evidence is far from conclusive.

In an Israeli cohort study of tenured working men, midlife BWV defined by three measurements of body weight was associated with higher risk of dementia 36 years later independent of the direction of weight changes[14]. Two nationwide cohort studies conducted in Korea also reported that BWV was related to a higher risk of Alzheimer’s and vascular dementia[15–16]. However, because of limited measurements of body weight, short-term follow-up, or both, it remains unclear whether the relation between BWV and dementia is causal or indicative of the early stage of dementia.

To overcome these limitations, we aimed to investigate the long-term association and temporal relation between BWV and the risk of dementia in the Health and Retirement Study (HRS). This prospective study provided a unique opportunity to evaluate repeated body weight measurements collected over 16 years before the regular assessment of dementia.

## 2. METHODS

### 2.1 Study population

This study was based on the Health and Retirement Study (HRS), a national longitudinal study among adults aged 50 years or older in the United States. From 1992, participants have been revisited every two years and asked about their basic status, sociodemographic, lifestyle and clinical information, etc. Before 2004, most follow-up interviews were conducted by telephone, except for respondents over the age of 80 who are offered face-to-face follow-up interviews. Since 2006, half of the samples were assigned a face-to-face interview with physical and biological measures and a psychosocial questionnaire. The other half completed only the core interview, usually by telephone[17]. A proxy respondent was sought for respondents who were unwilling or unable to complete an interview themselves due to health issues or death and were essential to maintaining coverage of the cognitively impaired to reduce attrition bias. About 9% of interviews were with a proxy respondent each wave, and 18% for those aged over 80. The response rate for HRS cohort was approximately 85%[17]. More detailed information can be found elsewhere (https://hrs.isr.umich.edu/).

Clinical-diagnosed dementia was self-reported on biennial follow-up questionnaires since 2008. We therefore chose the year 2008 as the study baseline and followed up participants thereafter until 2016 for incident dementia. The repeated body weight measures from 1992-2008 served as the long-term history of body weight. In this study, we included 5,547 participants in the HRS cohort (**Supplementary eFigure 1**) who: 1) completed at least 3 measurements of body weight during 1992-2008; 2) completed the baseline interview in 2008 and at least 1 interview thereafter; 3) were alive and free of dementia and other types of cognition- or memory-related diseases at baseline (in 2008).

### 2.2 Body weight variability

BWV was defined as the intraindividual instability and fluctuations in body weight during the process[11]. Participants reported their current body weight (kg) on biennial follow-up questionnaires from 1992 to 2008. We measured BWV using the coefficient of variation (CV) of body weight which was calculated as the quotient of standard deviation (SD) and mean over a period. We also measured BWV using standard deviation (SD) and variation independent of the mean (VIM) of sequential body weight assessments in the sensitivity analysis. Long-term (16-year) BWV was calculated as CV of body weight from 1992 to 2008.

### 2.3 Dementia and subtypes of dementia

The primary outcome in this study was all-cause dementia. The secondary outcomes were Alzheimer’s dementia and other types of dementia. The dementia status of participants was diagnosed by medical doctors or specialists such as neurologists and psychiatrists and reported by self- or proxy- respondents on biennial follow-up questionnaires in face-to-face, mailed, telephone, or internet-based surveys[18]. During the 8-year follow-up, 427 cases of incident dementia were reported, among which 201 were reported as Alzheimer’s dementia. To address the issue of potential underreporting of dementia, we calculated the total cognitive test score (0∼27 points) by summing the following cognitive items: immediate and delayed recall of a list of 10 words (1 point for each), five trials of serial 7s (i.e., subtract 7 from 100, and continue subtracting 7 from each subsequent number for a total of five trials, 1 point for each trial), and backward counting (2 points). We additionally identified participants with a cognitive function score <= 6 as dementia patients in sensitivity analysis, as is proposed in a previous study[19].

### 2.4 Other Covariates

Demographic and lifestyle factors, including age, gender, race (White/Caucasian / Black / Others), body weight (kg), height (m), education level (lower than high school / general educational development (GED) / high school / college / above), smoking status (never / former / current), alcohol consumption (never / former / current), household income (in quartiles), vigorous exercise frequency (>1 time per week / 1-3 times per month / never)[20], and medical status (hypertension, diabetes mellitus, cancer, heart disease, stroke, and other psychological diseases including emotional, nervous, and psychiatric problems) were collected using structured questionnaires biennially. These variables were considered as potential confounders and were adjusted in different models in thereafter analyses.

### 2.5 Statistical Analyses

Cox proportional hazard model was utilized to estimate the hazard ratio (HR) assessing the association between BWV and dementia. Survival time was calculated from baseline (2008) to incident dementia, death, dropout, or study endpoint (2016), whichever came first. In the primary analyses, long-term BWV was treated as both categorical (quartiles) and continuous variables. The multi-variable adjusted model included baseline age, gender, education level, household income, mean body weight, height, smoking status, alcohol consumption, vigorous exercise frequency, and long-term weight change (calculated as the end-to-end weight difference). The status of several major chronic morbidities was only included in the sensitivity analysis because they may act as mediators in the causal pathway of BWV and dementia. Single imputation was performed for missing values (4.6%) of these covariates, using mean for continuous variables and mode for categorical variables. And multiple imputations were performed in sensitivity analyses. Proportional hazard assumption was tested and verified using Schoenfeld’s residual methods and by including an interaction term with time in the model[21]. Tests for linear trend across BWV categories were calculated by entering BWV into Cox models as a continuous variable, and the potential non-linear association was tested using penalized smooth spline fit for BWV as a continuous variable. To examine the temporal relation between BWV and dementia, we analyzed the associations for successive 4-year BWVs assessed over time, with the period being divided into 1992-1996, 1996-2000, 2000-2004, and 2004-2008. We also estimated the adjusted HRs for the association of static body mass index (BMI) and its change from 1992 to 2008 with dementia.

In the secondary analyses, we further evaluated the association of BWV with subtypes of dementia, including Alzheimer’s dementia and other types of dementia. To test the potential effect modification of weight change, we categorized participants into weight loss (weight change < -5% compared to 1992), stable (-5% <= weight change <= 5% compared to 1992), and weight gain (weight change > 5% compared to 1992) [22–23]. Moreover, we conducted stratified analyses by major subgroups of age (≤70 vs. >70 years), gender, smoking status, alcohol consumption, hypertension, diabetes, stroke, psychological disease, and cancer. We evaluated the effect modifications by entering an interaction term of each variable and BWV in the Cox model.

In the post hoc sensitivity and exploratory analyses, we further adjusted the models with morbidity status (hypertension, diabetes mellitus, cancer, heart disease, stroke, and other psychological disease). To reduce the potential influence of varying number of body weight measures for different individuals, we conducted additional analyses only among participants with nine complete assessments of body weight over 16 years. For BWV measurements, we alternatively used SD and VIM in substitution for CV. To test the robustness of the temporal relationship, we mutually adjusted the BWVs in each period in the same model.

Statistical analyses were completed using SAS 9.4 (SAS Institute Inc, NC) and R 4.0.4. We reported 2-sided P values and 95% confidence intervals (CIs) throughout and P□<□0.05 was considered statistically significant.

## 3. RESULTS

### 3.1 Participants Characteristics

A total of 5,547 dementia-free participants were included in the study, among whom 56.7% were female, and 81.0% were White/Caucasian, and mean age at baseline was 71.1 years old (**Table 1**). During the follow-up period (mean = 6.8 y, median = 8 y), a total of 427 (7.7%) incident dementia cases were reported, among which 201 (47%) were Alzheimer’s dementia. Among dementia cases, the average time from baseline (2008) to dementia diagnosis was 5.1 y. Participants with the highest BWV were more likely to be female, have lower education levels (p<0.001), higher alcohol consumption (p<0.001), and less vigorous physical activity (p<0.001).

**Table 1.**
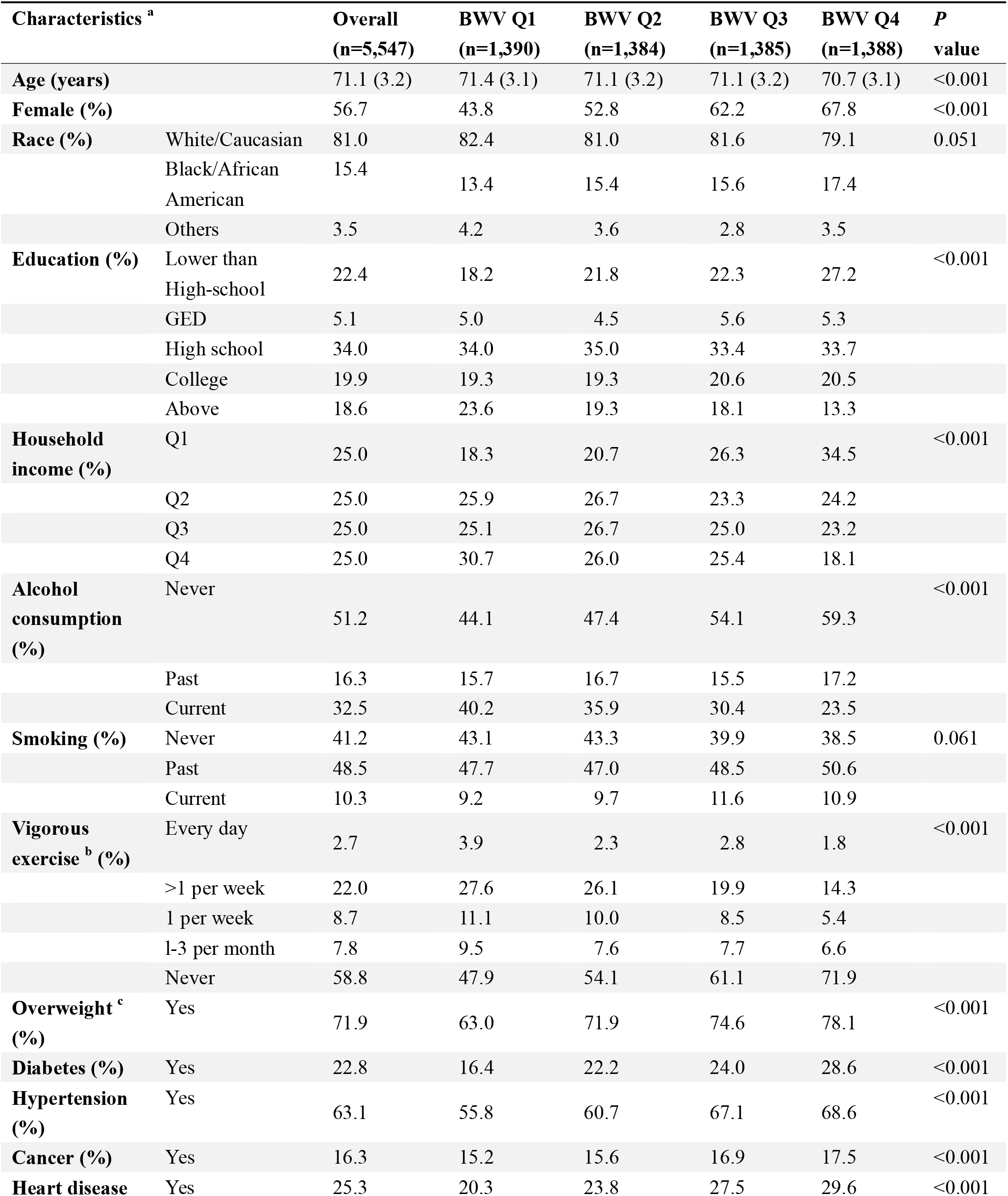
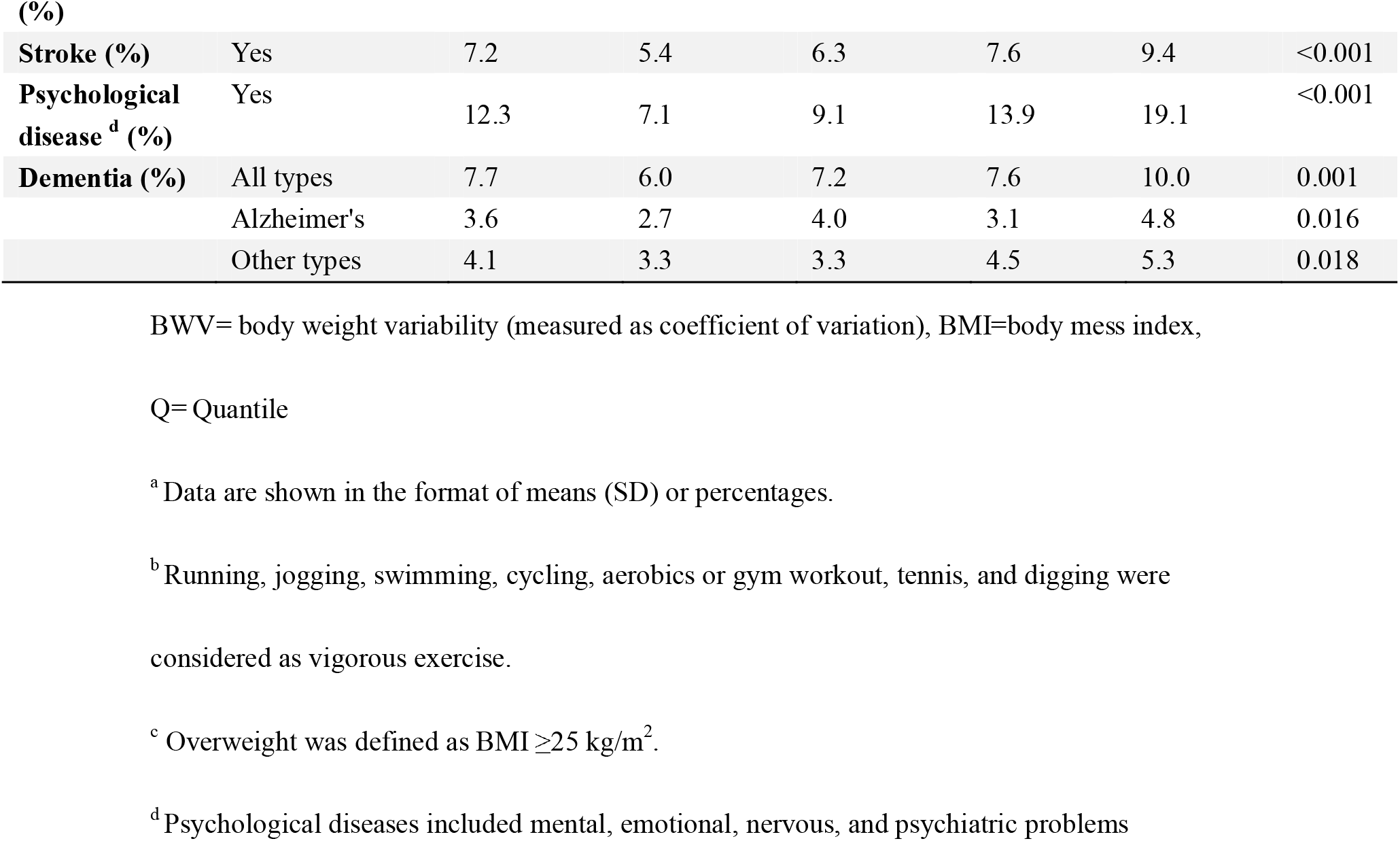
Baseline Characteristics of Participants According to Quartiles of BWV

### 3.2 Long-term body weight variability and dementia

In the multivariate-adjusted model (**Table 2**), participants with the highest quartile (Q4, median BWV=8.6%) of long-term BWV had a significantly higher risk of late-life incident dementia (HR=2.01, 95% CI, 1.48-2.72), compared to the bottom quartile (Q1, median BWV=2.5%). Each SD increment in BWV was associated with a 21% higher risk of dementia (HR=1.21, 95% CI,1.10-1.32, p-trend<0.001), with no significant non-linear relations observed for BWV and risk of dementia (p=0.81) (**Supplementary eFigure 2**). The independent associations of single-point BMI and successive 4-year body weight change with dementia were also presented in **Supplementary eFigure 3A & 3B**. After adjusting for major morbidities which potentially acted as mediators in the pathway of BWV to dementia, the association was only slightly attenuated (HR=1.18, 95%CI 1.07, 1.30). Similarly, the relation remained significant when using alternative BWV measurements such as SD (HR = 1.25, 95%CI 1.13, 1.39) and VIM (HR = 1.18, 95%CI 1.08, 1.29) (**Supplementary eTable 3).**

**Table 2.** Association of 16-years BWV with late-life onset of dementia (N=5,547)

|  | Median of<br>BWV (%) | Person-Time<br>(person-year) | Events | Model 1<sup>a</sup><br>HR (95% CI) | Model 2<sup>b</sup><br>HR (95% CI) | Model 3<sup>c</sup><br>HR (95% CI) |
| --- | --- | --- | --- | --- | --- | --- |
| **BWV (per SD)** | 4.6 | 37584 | 427 | 1.29 (1.18,1.40) | 1.21 (1.11,1.32) | 1.21 (1.10,1.32) |
| **BWV (Quantile)** |  |  |  |  |  |  |
| Q1 | 2.5 | 9778 | 83 | Reference | Reference | Reference |
| Q2 | 3.9 | 9526 | 100 | 1.32 (0.97, 1.80) | 1.27 (0.94, 1.73) | 1.31 (0.96, 1.79) |
| Q3 | 5.4 | 9380 | 105 | 1.43 (1.06, 1.94) | 1.34 (0.98, 1.82) | 1.37 (1.01, 1.87) |
| Q4 | 8.6 | 8990 | 139 | 2.24 (1.68, 2.99) | 1.92 (1.43, 2.58) | 2.01 (1.48, 2.72) |
| **P-trend** |  |  |  | <0.001 | <0.001 | <0.001 |

BWV=body weight variability (measured as coefficient of variation), SD=standard deviation,
HR=hazard ratio, CI=confidence interval
<sup>a</sup> Model 1: adjusting for age and gender.
<sup>b</sup> Model 2: further adjustment for race, education, household income, height, smoking status, alcohol, and exercise in addition to model 1.
<sup>c</sup> Model 3: further adjustment for mean body weight and body weight change over 16 years in addition to model 2.

### 3.3 Temporal relationship between body weight variability and dementia

We observed a significant association even for 4-year BWV calculated from 12 years to 8 years before study baseline (HR=1.13, 95% CI, 1.03-1.23, p-trend<0.001), i.e. on average 15 years prior to dementia diagnosis (**Figure 1A & Supplementary eTable 1**), especially among participants with minor long-term weight change (<= ±5%) (**Figure 1B**). The association became stronger when the 4-year BWV was assessed closer to study baseline. Each SD increment in the latest 4-year BWV was associated with a 19% increased risk of dementia (HR=1.19, 95% CI, 1.10-1.29, p-trend<0.001). The estimates remained similar after including BWVs of different periods into the same model (data not shown).

**Figure 1.**
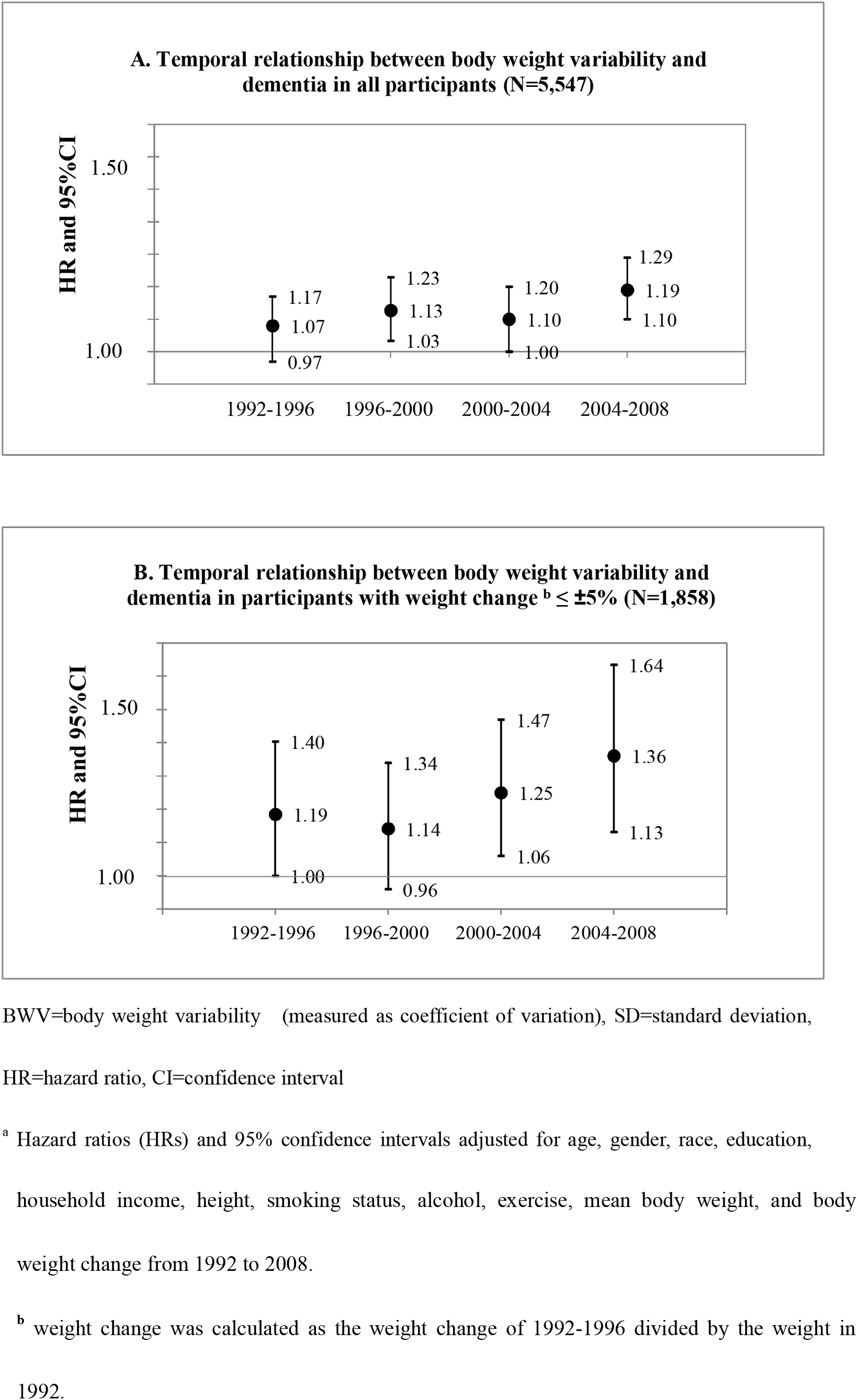
Temporal relations ^a^ between BWV (per SD increment) and dementia in all participants (A) and in participants with long-term weight change <5% (B) from 1992 to 2008.

### 3.4 Secondary and sensitivity analyses

For major subtypes of dementia, similar associations were observed for both Alzheimer’s dementia (HR_Q4vsQ1_ = 1.84, 95%CI 1.15, 2.92) and other types of dementia (HR_Q4vsQ1_ = 2.03, 95%CI 1.35, 3.04) (**Table 3**). Moreover, we observed consistent associations between BWV and dementia across most subgroups (e.g., age, gender, smoking status, etc.), although the association was stronger among participants with heart diseases (p-interaction=0.001), with stroke (p-interaction<0.001), and without historical psychological diseases (p-interaction=<0.001) (**Figure 2 & Supplementary eTable 2**). Alought no significant effect modification by weight change status was observed (p-interaction=0.435), the association was stronger among the subgroup of participants with minor long-term weight change (HR=1.45, 95%CI, 1.19, 1.79, p < 0.001). In sensitivity analyses, we observed robust associations of BWV with dementia. When only participants who completed all body weight measurements within 16 years were included, the association were generally the same as in primary analysis, with each SD increment in the long-term BWV associated with a 25% increased risk of dementia (HR = 1.25, 95%CI 1.13, 1.39). Detailed results of sensitivity analyses could be found in **Supplementary eTable 3**.

**Figure 2.**
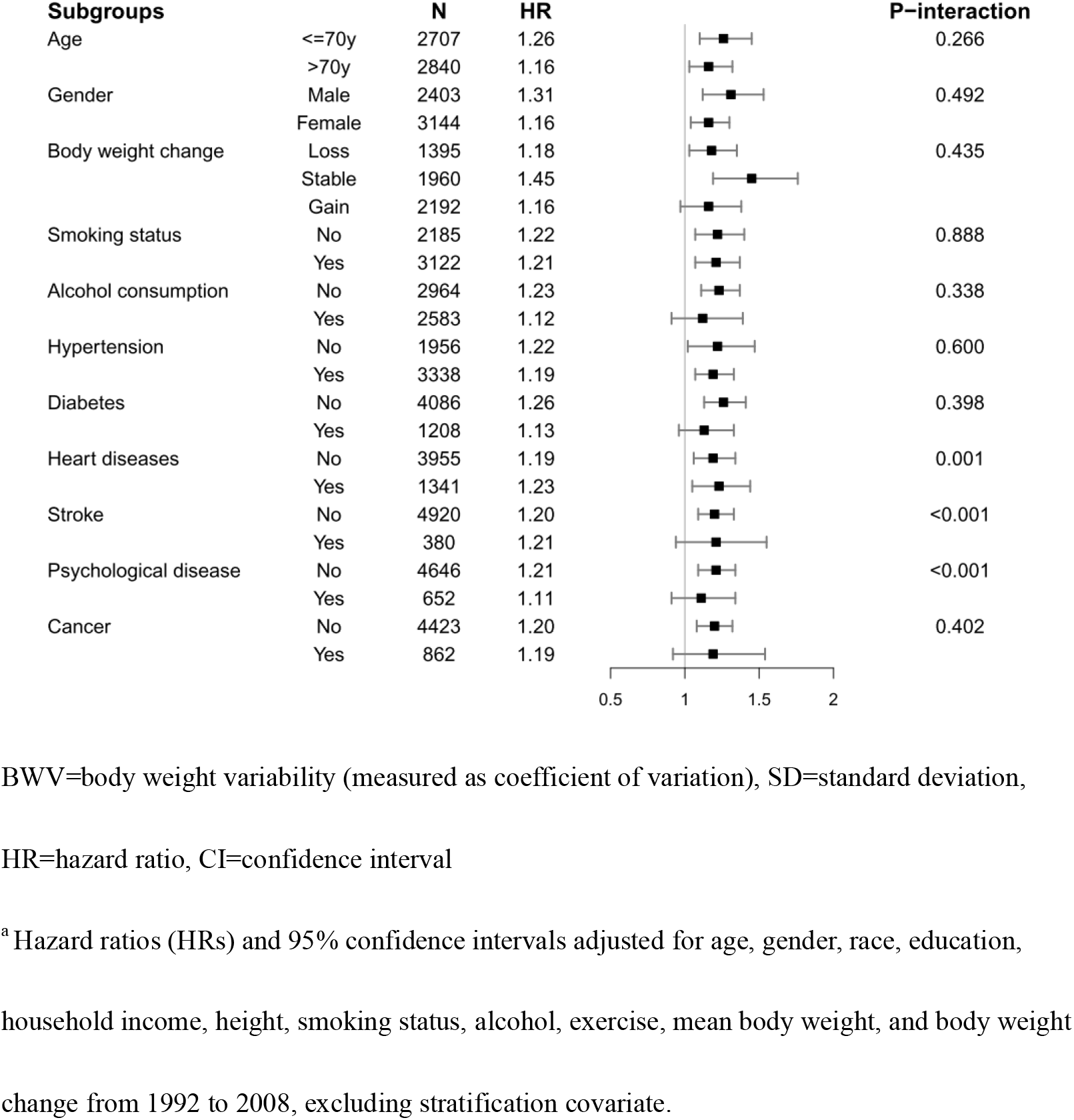
Association^a^ of 16-year BWV (per SD increment) with late-life onset of dementia in subgroups.

**Table 3.**
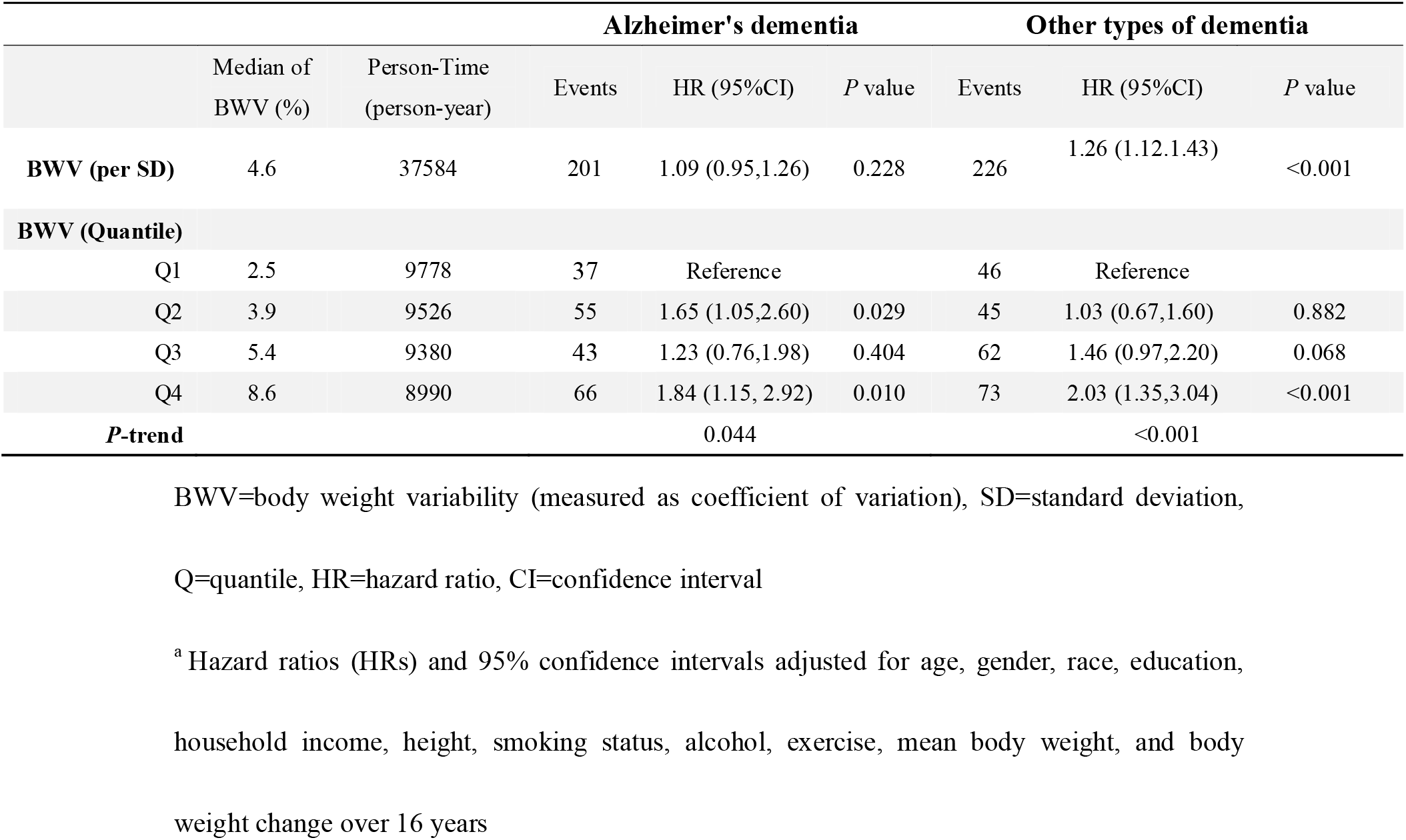
Association of 16-years BWV with late-life onset of dementia subtypes ^a^ (N=5,547)

## 4. DISCUSSION

In this prospective study of older adults in the U.S., greater long-term variability in body weight was associated with higher risk of dementia in later life beyond body weight and its change. This association was similar across major subgroups. This significant association was even observed for BWV estimated approximately 15 years before dementia diagnosis and became more pronounced when BWV was estimated proximal to diagnosis. In addition, the associations remained similar among a subgroup of participants with stable body weight level. In aggregate, these results strongly suggested that body weight variability may be a novel risk factor for dementia.

To our knowledge, three population-based studies have evaluated the relation between BWV and dementia, all reporting higher BWV being related to dementia[14–16]. Two nationwide studies conducted in Korean population respectively found that the highest BWV (calculated as VIM using repeated assessment of body weight) was associated with approximately 39%-42% higher risk of dementia onset in the follow-up period[15],[16], and the association was stronger for participants who were underweight. Another study conducted in Israel[14] also reported that higher BWV (SD2.66 kg vs. 1.15 kg) assessed 36 years before baseline was associated with a higher late-life dementia incidence (HR = 1.74, 95%CI 1.14∼2.64). Despite multiple differences in study designs and study populations, our results were similar to previous findings, providing strong additional evidence that larger long-term BWV was related to a higher risk of dementia. Moreover, we observed that higher BWV was associated with even increased risk of dementia among participants with stable weight.

The significant association for BWV measured approximately 15 years before dementia diagnosis indicated that BWV might have a long-term association with future dementia, which could also be collaboratively supported by the aforementioned study in Israel with a time window of more than three decades between BWV and the development of dementia. We also found that the corresponding association became stronger when BWV was measured closer to dementia diagnosis, suggesting that BWV could also be a precursor or early marker of dementia. Overall, BWV could act both as a risk factor and as an early sign of dementia, the entwined relation between the two warrants further research.

Although the underlying mechanism remains unclear and complex, several possible pathways could explain this association. Previous animal experimental studies have indicated that greater BWV over a long-term period may induce chronic neuroinflammatory response[24–26]. For example, weight-cycling increased the number of immune-related T cells in adipose tissue[27–28] and C-reactive protein level [29–30], which may subsequently lead to cognitive impairment and progression to dementia. BWV might also exert oblique influence on cognition function via unbalancing metabolic homeostasis[27, 31]. Furthermore, recent studies also found that weight-related microflora can interact with multiple nodes of the metabolic, endocrine, and immune systems through the gut-brain axis[32], and eventually lead to mental disorders and neurodegenerative diseases[33–35]. Since both BWV and neurodegenerative diseases are closely related to homeostasis and balance in the body, it is arbitrary to affirm the causal relationship based on current evidence, but to the least, our findings supported that BWV was strongly associated with the risk of subsequent dementia. To further reveal the complex causal relation and the underlying mechanism, more research, both experimental and observational, is needed in exploring this association.

The present study has several strengths. To our knowledge, our study is one of the few in reporting the association between long-term (up to 16-year) body weight variability and the risk of incident dementia and the first in temporal relation between the two. Moreover, the prospective study design, long-term follow-up, high follow-up rate, and careful control of various potential confounders minimized potential bias and provided relatively valid estimates of associations. This study made the best use of the repeated weight measures over 16 years to minimize possible measurement error and represent long-term BWV. Our findings should be interpreted with caution due to some limitations. First, since the outcome was reported by the participants themselves or proxy respondents, it was possible that the cases were underreported. However, it did not essentially undermine our conclusion: we included people with low cognitive function scores in the sensitivity analysis, which did not change the results. Second, it remains possible that residual confounding, or unmeasured confounding such as diet, exists as with other observational studies. In addition, the study population mainly consisted of Caucasians in the United States, which may limit the generalizability of our findings. Nevertheless, there is currently no prior finding suggesting that the relation would be qualitatively different across ethnicity.

## 5. CONCLUSIONS

Our prospective findings relating BWV over 16 years to the risk of dementia in later life provided further evidence that greater BWV may be a novel risk factor for dementia. Among individuals with only slight weight change, greater weight variability during the change process is associated with a significantly higher risk of dementia. Considering the irreversible nature of dementia, clinical and public health practice recommendations should pay attention to the management of body weight variability among older adults. Further study is needed to confirm the study findings across races and regions and study the underlying biological mechanisms.

## Data Availability

Data described in the manuscript and the codebook will be made available upon request pending application and approval (https://hrs.isr.umich.edu/).

## ACKNOWLEDGEMENT

C. Y. and Y. M. designed the study; H. C. and T. Z. performed the statistical analyses; C., and Y. M. interpreted the data; H. C., T. Z. draft the manuscript, W. X., J. J., L. H., J. G., G. Z., X. L., Y. Z., and A. H. further revised the manuscript; C. Y. supervised the data analysis and interpretation; C.Y. had the primary responsibility for the study final content. All authors critically reviewed the manuscript and approved the final draft.

## FUNDING

This study was funded by the Zhejiang University Education Foundation Global Partnership Fund. The Health and Retirement Study is sponsored by the National Institute on Aging (NIA U01AG009740) and the Social Security Administration. The study director is Dr. David R. Weir of the Survey Research Center at the University of Michigan’s Institute for Social Research. The funding organizations had no role in the design and conduct of the study; collection, management, analysis, and interpretation of the data; preparation, review, or approval of the manuscript; and decision to submit the manuscript for publication.

## DECLARATIONS OF INTEREST

None.

## Abbreviations

(HRS): Health and Retirement Study
(BWV): body weight variability
(HR): hazard ratio
(CI): confidence interval
(SD): standard deviation
(CV): coefficient of variation
(VIM): variation independent of mean
(BMI): body mass index

## Notes

This work was supported by the Zhejiang University Education Foundation Global Partnership Fund. The funding agency had no role in study design, data collection, analysis, decision to publish, or manuscript preparation.

### Competing Interest Statement

The authors have declared no competing interest.

### Author Declarations

UM Health Sciences/Behavioral Sciences IRB Protocol: HUM00061128 Approved through 10/18/2018 Associated protocols: HUM00056464, HUM00002562, HUM00074501, HUM00079949, HUM00080925, HUM00085942, HUM00099822, HUM00103072, HUM00106904, HUM00122335, REP00000046

