## Supplementary Files for "Association of long-term body weight variability with dementia: a prospective study"

### Supplementary Online Content

**Supplementary Figure Legends**

**eFigure 1.** Inclusion and Exclusion criteria

**eFigure 2.** Body weight variability and all-cause dementia risk

**eFigure 3.** Hazard ratios (HRs) of dementia risk associated with body mass index (BMI) and it change from 1992 to 2008

**Supplementary Online Tables**

**eTable 1.** Temporal relations between body weight variability and dementia in all participants.

**eTable 2.**  Association of 16-year body weight variability with late-life onset of all-cause dementia in different sub-groups.

**eTable 3.**  Sensitivity analyses of association of 16-years body weight variability (BWV) with late-life onset of dementia.

**eFigure 1. Inclusion and Exclusion criteria**

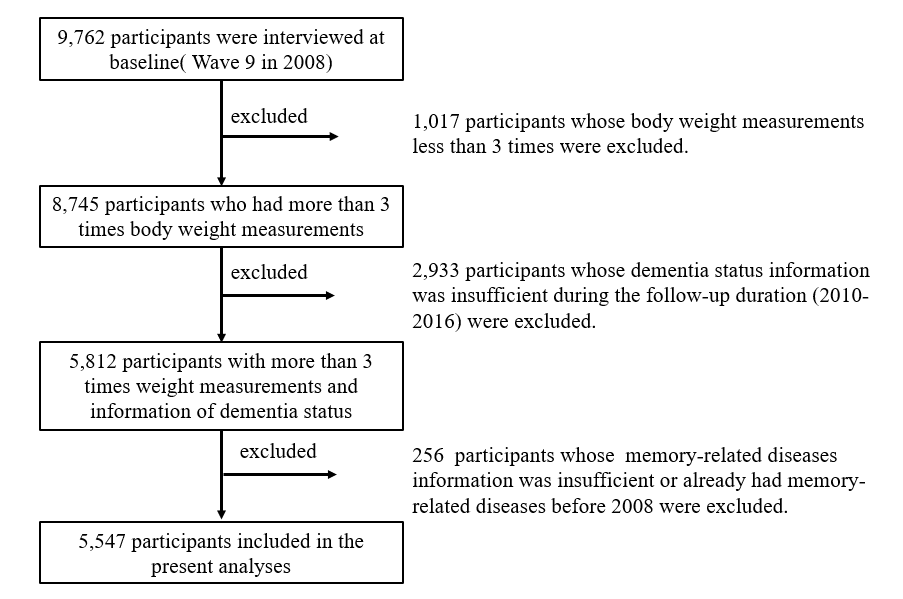

**eFigure 2. Body weight variability and all-cause dementia risk^a^**

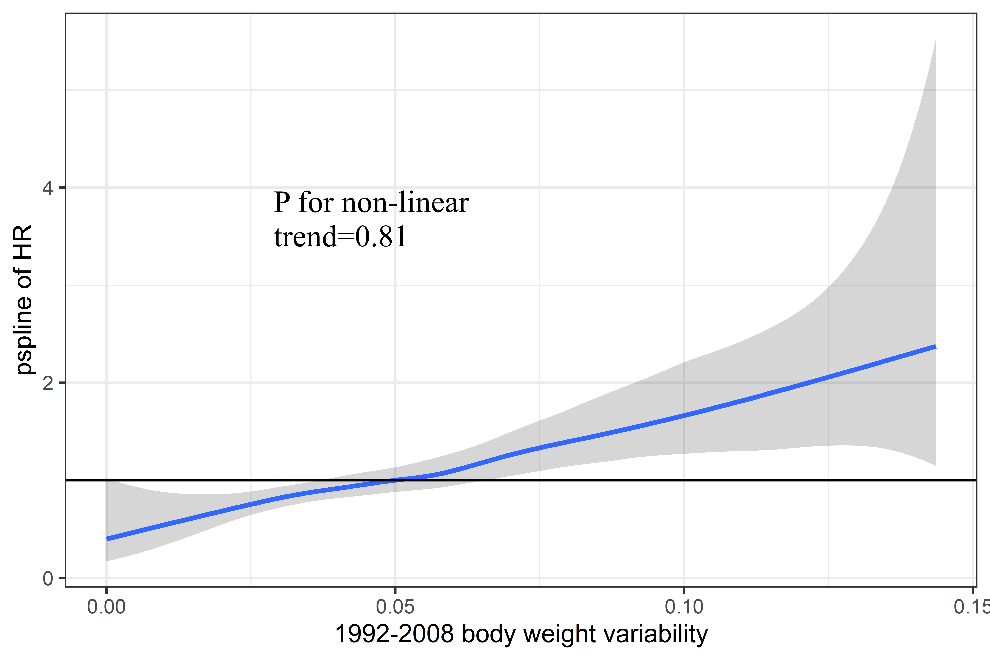

^a^ Adjusted for age, gender, race, education, household income, height, smoking status, alcohol, exercise, mean body weight, and body weight change from 1992 to 2008.

**eFigure 3. Hazard ratios (HRs) ^a^ of dementia risk associated with body mass index (BMI) and body weight change rate from 1992 to 2008**

A: Body mass indexb (per 1kg/m2) from 1992 to 2008 and dementia in late life  (N=5,547)

|  |  |  | 1992 1994 1996 1998 2000 2002 2004 2006 2008 | Moderate SCF | Poor SCF |
| --- | --- | --- | --- | --- | --- |
|  |  |  |  | -0.5 | -1 |
| 1992 | 1.031 | 0.988 | 1.009 |  |  |
| 1994 | 1.035 | 0.991 | 1.013 |  |  |
| 1996 | 1.029 | 0.986 | 1.008 |  |  |
| 1998 | 1.03 | 0.988 | 1.009 |  |  |
| 2000 | 1.024 | 0.982 | 1.003 |  |  |
| 2002 | 1.014 | 0.972 | 0.993 |  |  |
| 2004 | 1.005 | 0.964 | 0.984 |  |  |
| 2006 | 0.993 | 0.952 | 0.972 |  |  |
| 2008 | 0.979 | 0.939 | 0.959 |  |  |

B: Body weight change (per percentage)c from 1992 to 2008 and dementia in late life (N=5,547)

|  | 1992-2008 1992-1996 1996-2000 2000-2004 2004-2008 |  |  | Moderate SCF | Poor SCF |
| --- | --- | --- | --- | --- | --- |
|  |  |  |  | -1 | -1 |
| 1992-2008 | 1.02 | 1 | 1.009 |  |  |
| 1992-1996 | 1.026 | 1 | 1.01 |  |  |
| 1996-2000 | 1.035 | 1 | 1.018 |  |  |
| 2000-2004 | 1.03 | 1 | 1.012 |  |  |
| 2004-2008 | 1.047 | 1 | 1.033 |  |  |

CV=coefficient variation, SD=standard deviation, BMI=body mass index, HR=hazard ratio, CI=confidence interval

^a^ HR of the highest quartile (Q4) vs. the lowest quartile (Q1) of BMI or its absolute change (per percentage) adjusted for age, gender, race, education, household income, height, smoking status, alcohol, exercise, mean body weight, and body weight change from 1992 to 2008.

^b^ BMI was calculated as weight in kilograms divided by the square of height in meters.

^c^ Absolute body weight change (percentage) was calculated as the absolute change of body weight within 4 years divided by the weight measured at the start of the 4 years.

**eTable 1. Temporal relations between body weight variability and dementia in all participants.**

|  | | | | Model 1^1^ | | Model 2^2^ | | Model 3^3^ | |
| --- | --- | --- | --- | --- | --- | --- | --- | --- | --- |
|  | | | Median (%) | HR (95%CI) | *P* | HR (95%CI) | *P* | HR (95%CI) | *P* |
| 1992-1996 |  | |  | | | | | | |
| CV (per SD) |  | | 2.8 | 1.13 (1.04, 1.23) | 0.004 | 1.08 (0.99, 1.18) | 0.066 | 1.07 (0.97,1.17) | 0.163 |
| CV (Quantile) |  | |  | | | | | | |
| 1 | | 0.8 | | ref | ref | ref | ref | ref | ref |
| 2 | | 2.1 | | 0.96 (0.72, 1,27) | 0.751 | 0.97 (0.73, 1.30) | 0.850 | 0.95 (0.71, 1.29) | 0.764 |
| 3 | | 3.6 | | 1.10 (0.83, 1.46) | 0.408 | 1.05 (0.79, 1.40) | 0.104 | 1.07 (0.79, 1.44) | 0.658 |
| 4 | | 6.5 | | 1.42 (1.08, 1.87) | 0.012 | 1.30 (0.97, 1.72) | 0.063 | 1.28 (0.96, 1.72) | 0.094 |
| *P*-trend | |  | | 0.007 | | 0.049 | | 0.063 | |
| 1996-2000 |  | |  | | | | | | |
| CV (per SD) |  | | 2.8 | 1.15(1.06, 1.25) | 0.001 | 1.12(1.02, 1.21) | 0.014 | 1.13(1.03, 1.23) | 0.007 |
| CV (Quantile) |  | |  | | | | | | |
| 1 | | 0.8 | | ref | ref | ref | ref | ref | ref |
| 2 | | 2.1 | | 0.96 (0.72, 1.29) | 0.800 | 0.96(0.72, 1.28) | 0.762 | 1.03(0.75,1.42) | 0.850 |
| 3 | | 3.6 | | 1.10 (0.83, 1.46) | 0.436 | 1.09(0.82, 1.44) | 0.550 | 1.21(0.89,1.64) | 0.234 |
| 4 | | 6.5 | | 1.56 (1.19, 2.03) | 0.001 | 1.42(1.09, 1.85) | 0.010 | 1.59(1.18,2.14) | 0.002 |
| *P* -trend | |  | | 0.001 | | 0.007 | | 0.001 | |
| 2000-2004 |  | |  | | | | | | |
| CV (per SD) |  | | 2.9 | 1.17(1.07, 1.28) | <.0001 | 1.16(1.02, 1.22) | 0.015 | 1.10(1.00, 1.20) | 0.048 |
| CV (Quantile) |  | |  | | | | | | |
| 1 | | 0.8 | | ref | ref | ref | ref | ref | ref |
| 2 | | 2.2 | | 1.08 (0.81, 1.45) | 0.580 | 1.09(0.82, 1.46) | 0.567 | 1.30(0.94, 1.78) | 0.107 |
| 3 | | 3.6 | | 1.21 (0.91, 1.60) | 0.197 | 1.17 (0.88, 1.55) | 0.298 | 1.32(0.96, 1.82) | 0.087 |
| 4 | | 6.7 | | 1.51 (1.14, 1.98) | 0.003 | 1.37 (1.04, 1.80) | 0.027 | 1.61(1.18, 2.20) | 0.003 |
| *P* -trend | |  | | 0.003 | | 0.023 | | 0.004 | |
| 2004-2008 |  | |  | | | | | | |
| CV (per SD) |  | | 2.9 | 1.28(1.20, 1.37) | <0.001 | 1.22(1.14, 1.31) | <0.001 | 1.19 (1.10,1.29) | <0.001 |
| CV (Quantile) |  | |  | | | | | | |
| 1 | | 0.9 | | ref | ref | ref | ref | ref | ref |
| 2 | | 2.2 | | 0.94 (0.69,1.28) | 0.695 | 0.94 (0.69, 1.29) | 0.700 | 1.03(0.74, 1.44) | 0.875 |
| 3 | | 3.6 | | 1.30 (0.98,1.74) | 0.072 | 1.27(0.95, 1.69) | 0.112 | 1.42(1.04, 1.93) | 0.027 |
| 4 | | 6.7 | | 1.81 (1.38,2.38) | <0.001 | 1.63 (1.24, 2.15) | <0.001 | 1.62(1.20, 2.21) | 0.002 |
| *P* -trend | |  | | <0.001 | | <0.001 | | <0.001 | |

CV=coefficient variation, SD= standard deviation, HR=hazard ratio, CI=confidence interval

^1^Model 1: adjusted for age and gender.

^2^Model 2: model 1 + race, education, household income, height, smoking status, alcohol, and exercise.

^3^Model 3: model 2 + mean body weight and body weight change over 16 years.

**eTable 2. Association of 16-year body weight variability with late-life onset of all-cause dementia in different sub-groups.**

| Subgroups of HRS | | HR ^a^ (95% CI) vs Q1 | | | |
| --- | --- | --- | --- | --- | --- |
|  |  | CV (per SD) | Q2 | Q3 | Q4 |
| Age | ≤70y | 1.26 (1.10,1.45) ** | 1.15 (0.61,2.17) | 1.48 (0.80,2.73) | 2.77 (1.56,4.93) * |
|  | >70y | 1.16 (1.03,1.32) * | 1.40 (0.98,2.00) | 1.33 (0.93,1.91) | 1.68 (1.16,2.43) ** |
| Gender | Male | 1.31 (1.12,1.53) ** | 1.20 (0.76, 1.89) | 1.52 (0.96,2.38) | 1.87 (1.16,3.00) ** |
|  | Female | 1.16 (1.04,1.30) * | 1.36(0.89,2.10) | 1.26(0.82,1.93) | 2.05(1.37,3.08) ** |
| Body weight change ^b^ | Loss | 1.18 (1.03,1.35) * | 0.86(0.40,1.84) | 1.13(0.56,2.29) | 1.56(0.75,3.23) |
|  | Stable | 1.45 (1.19,1.76) ** | 1.79(1.20,2.67) ** | 1.40 (0.84,2.32) | 2.28(1.31,4.00) ** |
|  | Gain | 1.16 (0.97,1.38) | 1.03(0.50,2.15) | 1.30(0.64,2.63) | 1.81(0.86,3.79) |
| Smoking | No | 1.22 (1.07,1.40) ** | 1.65(1.03,2.64) * | 1.60(0.99,2.58) | 2.46(1.54,3.91) ** |
|  | Yes | 1.21 (1.07,1.37) ** | 1.12(0.73,1.70) | 1.24(0.82,1.86) | 1.77(1.18,2.64) ** |
| Drinking | No | 1.23 (1.11,1.37) ** | 1.24(0.83,1.86) | 1.32(0.90,1.96) | 2.20(1.52,3.20) ** |
|  | Yes | 1.12 (0.91,1.39) | 1.41(0.86,2.31) | 1.45(0.87,2.43) | 1.62(0.93,2.82) |
| Hypertension | No | 1.22 (1.02,1.47) * | 1.52(0.92,2.51) | 1.37(0.81,2.33) | 1.46(0.84,2.56) |
|  | Yes | 1.19 (1.07,1.33) ** | 1.21(0.81,1.82) | 1.38(0.94,2.02) | 2.17(1.50,3.15) ** |
| Diabetes | No | 1.26 (1.13,1.41) ** | 1.43(1.00,2.05) * | 1.31(0.91,1.89) | 2.15(1.51,3.07) ** |
|  | Yes | 1.13 (0.96,1.33) | 1.02(0.53,1.94) | 1.51(0.83,2.73) | 1.80(0.99,3.26) |
| Heart disease | No | 1.19 (1.06,1.34) ** | 1.44(1.00,2.06) * | 1.38(0.95,1.99) | 1.86(1.28,2.69) ** |
|  | Yes | 1.23 (1.05,1.44) * | 1.05(0.56,1.94) | 1.28(0.72,2.28) | 2.12(1.21,3.71) ** |
| Stroke | No | 1.20 (1.09,1.33) ** | 1.28(0.92,1.79) | 1.29(0.93,1.80) | 1.98(1.44,2.75) ** |
|  | Yes | 1.21 (0.94,1.55) | 1.50(0.53,4.21) | 2.18(0.81,5.91) | 2.20(0.84,5.78) |
| Psychological disease ^c^ | No | 1.21 (1.09,1.34) ** | 1.62(1.16,2.28) ** | 1.48(1.04,2.11) * | 2.25(1.59,3.19) |
|  | Yes | 1.11 (0.91,1.34) | 0.34(0.13,0.83) * | 0.69(0.36,1.34) | 0.90(0.48,1.69) |
| Cancer | No | 1.20 (1.08,1.32) ** | 1.29(0.92,1.82) | 1.46(1.05,2.04) * | 1.95(1.40,2.72) ** |
|  | Yes | 1.19 (0.92,1.54) | 1.29(0.58,2.90) | 0.94(0.39,2.22) | 2.05(0.92,4.53) |

* *P* <0.05, ** *P* <0.01

CV=coefficient variation, Q=quantile, SD=standard deviation, BMI=body mass index, HR=hazard ratio, CI=confidence interval

^a^ HRs and 95% CIs adjusted for age, gender, race, education, household income, height, smoking status, alcohol, exercise, mean body weight, and body weight change over 16 years.

^b^ body weight change: weight loss (weight change < -5% of baseline weight), stable (-5% <= weight change <= 5% of baseline weight), and weight gain (weight change > 5% of baseline weight)

^c^ Psychological disease including emotional, nervous, and psychiatric problems

**eTable 3. Sensitivity analyses of association of 16-years body weight variability (BWV) with late-life onset of dementia.**

|  | Median of BWV | ^a^ HR (95% CI) | p-trend |
| --- | --- | --- | --- |
| Only including participants who completed all measurements of body weight before baseline (N=4,484). | | | |
| BWV (per SD increment) | 4.5% | 1.25(1.13,1.39) | <0.001 |
| BWV calculated as standard deviation (SD) | | | |
| BWV (per SD increment) | 3.55 | 1.25 (1.13, 1.39) | <0.001 |
| BWV calculated as variability independent of mean ^b^ (VIM) | | | |
| BWV (per SD increment) | 0.45 | 1.18 (1.08, 1.29) | <0.001 |
| Dementia defined as cognition score ^c^ ≤6. | | | |
| BWV (per SD increment) | 4.6% | 1.22(1.14,1.31) | <0.001 |
| Morbidities ^d^ adjusted | | | |
| BWV (per SD increment) | 4.6% | 1.18(1.07,1.30) | <0.001 |
| Adjusting body weight change calculated as participant-specific slope of linear regression | | | |
| BWV (per SD increment) | 4.6% | 1.20(1.10,1.31) | <0.001 |

BWV=body weight variability, SD=standard deviation, VIM=variation independent of mean, HR=hazard ratio, CI=confidence interval

**^a^** Adjusted for Age, gender, race, education, household income, height, smoking status, alcohol, exercise, mean body weight and body weight change over 16 years

^b^ VIM: calculated as the SD divided by the mean to the power x and multiplied by the population mean to the power x. The power x is obtained by fitting a curve through a plot of SD against mean using the model SD = 100*mean^x^.

^c^ Sum of the following cognitive items: immediate and delayed recall of a list of 10 words (1 point for each), five trials of serial 7s (i.e., subtract 7 from 100, and continue subtracting 7 from each subsequent number for a total of five trials, 1 point for each trial), and backward counting (2 points). We identified participants with a cognitive function score <= 6 as dementia patients.

^d^ Morbidities include hypertension, diabetes mellitus, cancer, heart disease, stroke, and other psychological diseases including emotional, nervous, and psychiatric problems.
